# Incorporating Angina into the H_2_FPEF Score Improves Diagnostic Performance for HFpEF in Women

**DOI:** 10.1101/2025.10.30.25339191

**Authors:** Kaiyong Qu, N. Charlotte Onland-Moret, Amber de Vos, Anne-Mar van Ommen, Yvonne van Mourik, Evelien E. van Riet, Leandra J.M. Boonman-Winter, Marc L. Handoko, Maarten J. Cramer, Arco J. Teske, Roxana Menken, Frans H. Rutten, Hester M. den Ruijter, Elisa Dal Canto

## Abstract

**Background and Aims:** The H_2_FPEF score is a widely used prediction tool used during the diagnostic work-up of heart failure with preserved ejection fraction (HFpEF). However, having angina symptoms is not included in the score, despite being common in patients with HFpEF. We hypothesize that incorporating angina in the H_2_FPEF score may improve its performance. Given the known sex differences in HFpEF, sex-specific analyses are warranted.

**Methods:** We included 1,266 individuals suspected HFpEF, with 515 from the UHFO-DM cohort and 751 from a combination cohort of STRETCH, TREE and UHFO-COPD. Participants underwent standardized symptom collection, including angina, using WHO questionnaires and expert-panel adjudication of HFpEF. Following evaluation of H_2_FPEF, we assessed the association of angina with HFpEF independent of H_2_FPEF using logistic regression. By adding angina to H_2_FPEF, we developed a modified algorithm and evaluated it by AUC, calibration, reclassification, and decision curve analysis. All analyses were stratified by sex.

**Results:** In the UHFO-DM cohort, HFpEF prevalence was 24%. Overall H_2_FPEF discrimination (AUC) was 0.72, with 0.69 in women and 0.74 in men. Angina was independently associated with HFpEF in women (OR 3.96, 95% CI 1.72–9.11, P=0.001) but not in men (1.90, 0.88–4.10, 0.102). This was also found in the combination cohort (women: 2.13, 1.14–3.97, 0.018; men: 0.85, 0.44–1.66, 0.638). In the UHFO-DM cohort, adding one point for angina in a modified H_2_FPEF score in women improved AUC from 0.69 to 0.71 (DeLong P=0.030), increased sensitivity (0.53 to 0.60) and negative predictive value (0.80 to 0.82), and yielded a continuous net reclassification improvement of 0.449, with preserved calibration and higher net clinical benefit on decision curves. No performance gain was observed with the same modification in men.

**Conclusions:** In women with suspected HFpEF, presence of angina provides diagnostic information independent of H_2_FPEF to uncover HFpEF. A simple sex-specific modification of H_2_FPEF, adding one point for angina in women, may slightly improve discrimination and rule-out performance in women.

**GRAPHICAL ABSTRACT:** 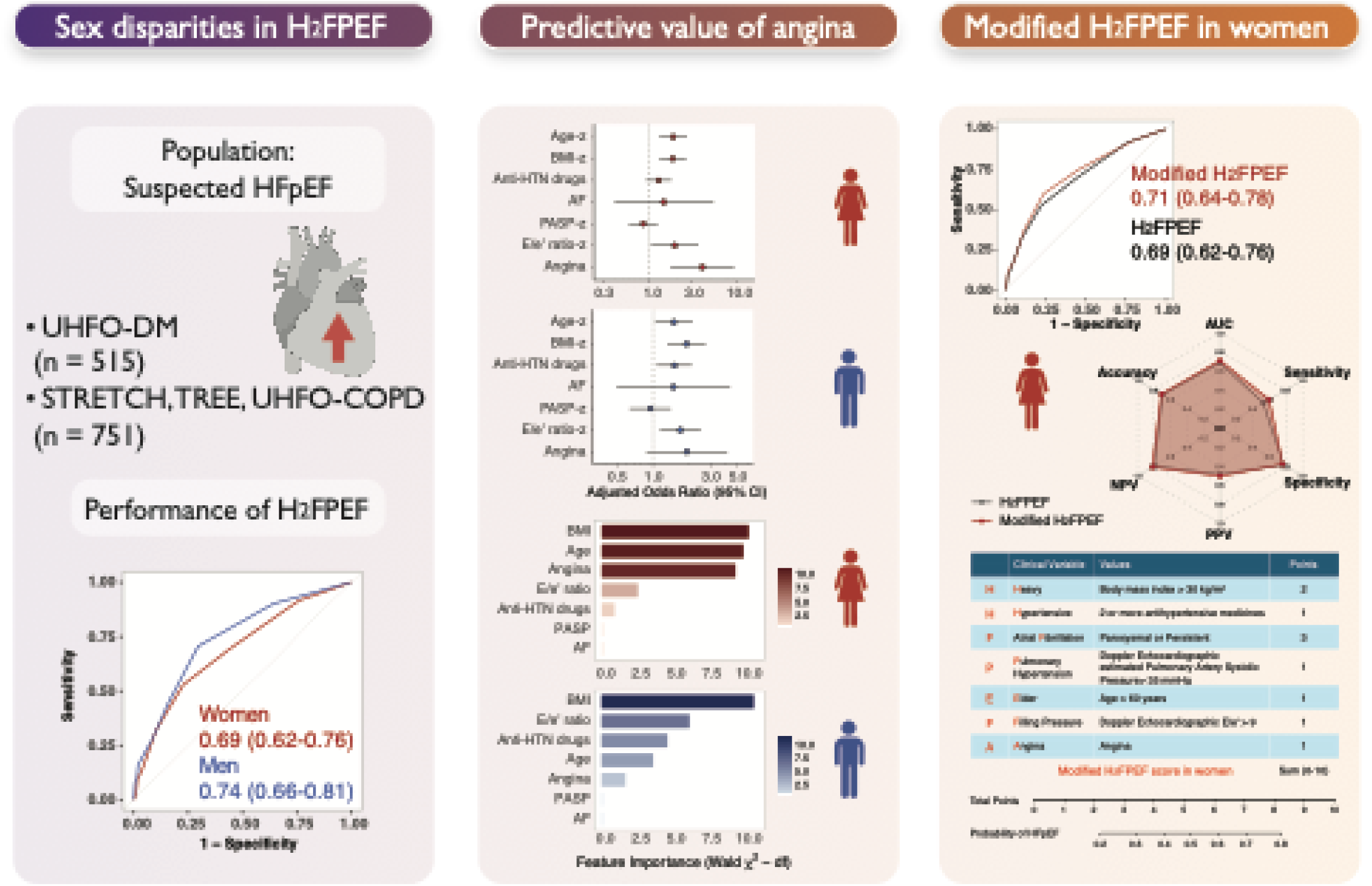

## Introduction

Heart failure with preserved ejection fraction (HFpEF) has emerged as a major global health concern, now accounting for approximately half of all heart failure cases(1). Its prevalence and incidence are steadily rising in parallel with aging populations and the increasing burden of risk factors such as hypertension, obesity, and diabetes(2). Patients with HFpEF experience high morbidity and mortality: one-year all-cause mortality rates approach 20–30% and 30-day readmission rates exceed 20% in contemporary cohorts(3). However, diagnosing HFpEF in practice is often challenging, notably in early stages in primary care(4). The reference standard for confirming HFpEF consists of invasive hemodynamic assessment demonstrating elevated filling pressures at rest or with exercise. It is impractical for routine evaluation due to its cost, risk of complications, and limited availability(5). Over the past few years, two major scoring systems have been proposed and adopted to aid HFpEF diagnosis. Current guidelines and expert consensus recommend the use of either the H_2_FPEF score or the HFA-PEFF algorithm as systematic approaches to estimate the probability of HFpEF(6, 7). The H_2_FPEF score (Heavy, Hypertensive, Atrial fibrillation, Pulmonary hypertension, Elder, Filling pressure) was derived in 2018 as a simple, evidence-based tool to distinguish HFpEF from non-cardiac causes of dyspnea(8). In contrast, the Heart Failure Association Pre-test assessment, Echocardiographic and biomarker evaluation, Functional test, Final etiologic workup (HFA-PEFF) algorithm is a more elaborate, stepwise scoring system endorsed by the European Society of Cardiology(9). Both the H_2_FPEF and HFA-PEFF algorithms have provided greater objectivity in HFpEF diagnosis, with the H_2_FPEF score characterized by simplicity and practicality in clinical practice. Nevertheless, important knowledge gaps remain. HFpEF is a markedly heterogeneous condition with pronounced sex-based differences. In community cohorts, women have approximately a 2:1 higher incidence of HFpEF compared with men(10). Moreover, women with HFpEF present more diastolic dysfunction (53% vs. 32%) and experience higher risks of all-cause mortality and heart failure readmission than men (HR=1.54)(11). Given these sex disparities, a sex-specific assessment of H_2_FPEF is warranted to ensure this algorithm is robust and equitable for both sexes. However, to the best of our knowledge, no study has yet evaluated the diagnostic accuracy of H_2_FPEF by sex. Another underexplored factor in HFpEF diagnosis is the role of angina pectoris. In the Duke Databank of 3,517 patients, about 40% of HFpEF patients reported angina symptoms(12). However, angina is not currently included in the H_2_FPEF score. The high frequency of angina in HFpEF patients may be explained by underlying obstructive coronary artery disease (CAD) and/or non-obstructive coronary artery disease (coronary vasospasm or coronary microvascular dysfunction (CMD)). Importantly, the population distribution of obstructive and non-obstructive CAD differs by sex. Non-obstructive CAD is more common in women and obstructive CAD is more common in men(13), which may influence how angina presents in HFpEF patients. Taken together, although angina is highly prevalent in HFpEF, it remains unknown whether it provides incremental diagnostic value beyond the H_2_FPEF score. In addition, given the known sex differences in HFpEF, sex-stratified evaluation of diagnostic tools is warranted. Considering these gaps, we designed the present study, to assess the sex-stratified diagnostic performance of the H_2_FPEF score, and to evaluate the association between angina and HFpEF independent of H_2_FPEF by sex. Furthermore, we sought to test whether incorporating angina into the H_2_FPEF algorithm improves its diagnostic performance.

## METHODS

### Population

This study is a cross-sectional analysis of patient data obtained from four independent cohorts conducted in primary care settings among high-risk individuals: the UHFO-DM, UHFO-COPD, STRETCH, and TREE cohorts(14–18). All cohorts shared the common objective to screen for previously unrecognized heart failure (HF) of any type. Data collection in all studies was performed using a standardized case record form that included questions on symptoms, medication use, medical history, physical examination findings, and results from additional investigations including electrocardiography, laboratory testing, and echocardiography. The UHFO-DM cohort (2009–2010) enrolled patients aged ≥60 years with type 2 diabetes and was used as the derivation cohort. The UHFO-COPD (patients labeled with COPD), STRETCH (patients who visited their general practitioner for shortness of breath), and TREE (Frail older people with short of breath) cohorts (2001–2012) included patients aged ≥65 years with shortness of breath as a common main symptom. These three cohorts were combined and used for external validation, as they contained more missing variables than UHFO-DM, despite their larger overall sample size. The primary endpoint was HFpEF, adjudicated by an expert panel of two cardiologists or one cardiologist and one pulmonologist together with an experienced general practitioner, based on all available diagnostic data (NT-proBNP was available only in TREE). Reproducibility of this method was high (κ = 0.84 from 10% re-evaluation). Patients without tissue Doppler imaging, with isolated right heart failure, left ventricular ejection fraction (LVEF) <50%, or adjudicated HFrEF were excluded. The study flow is shown in **Supplementary** Figure 1. All four studies were approved by the Medical Ethics Committee of the University Medical Center Utrecht, the Netherlands. All participants provided written informed consent.

### Baseline assessment

The presence of angina was assessed using the WHO questionnaires by a trained physician in a standardized manner. Hypertension, chronic obstructive pulmonary disease (COPD), asthma, valvular heart disease, cerebrovascular disease, and a history of myocardial infarction (MI), percutaneous coronary intervention (PCI), and coronary artery bypass grafting (CABG) were collected by self-report. Atrial fibrillation was determined by self-report and ECG. Dyslipidemia was defined as treatment for hypercholesterolemia. Patients were asked to bring their medication packages so that current drug treatment could be verified. Antihypertensive drugs included angiotensin-converting enzyme inhibitor/ angiotensin receptor blocker (ACEI/ARB), beta-blockers, calcium channel blocker (CCB), and diuretics. Blood samples were taken within two weeks of the diagnostic assessment for measurement of serum NT-proBNP and creatinine. Estimated glomerular filtration rate (eGFR) was calculated according to the CKD-EPI formula. Echocardiography was performed using a General Electric Vivid 7 imaging system (GE Vingmed Ultrasound AS, Horten, Norway) by well-trained and experienced cardiac sonographers. Details on the echocardiographic measurements are provided in the Supplementary Method.

### Statistical analysis

Baseline characteristics of the study population were stratified by sex and HFpEF diagnosis. Continuous variables were summarized as mean ± SD or median (IQR), and categorical variables as counts and percentages. Between-group comparisons were performed using Student’s t-test or Mann–Whitney U test for continuous variables and χ² or Fisher’s exact test for categorical variables, as appropriate. Missing data (**Supplementary Table 1**) were handled using multiple imputation by chained equations (MICE), with 10 imputations and 10 iterations per imputation. Imputations were performed separately by sex and cohort. In the UHFO-DM cohort, logistic regression analyses were conducted to examine the association between determinants and HFpEF. Univariable analyses were first performed for angina and each of the six individual variables comprising the H_2_FPEF score. Multivariable models were then constructed to assess whether angina provided additional predictive value beyond the individual H_2_FPEF components. Subsequently, the total H_2_FPEF score was taken as a single variable with angina in multivariable models. All models were stratified by sex. In the combined validation cohort, the same analytical strategy was applied. However, due to the high rate of missing data for pulmonary artery systolic pressure (PASP) (∼90%), this variable was excluded from models, and the H_2_FPEF score was therefore not computed. Variable importance was evaluated by the Wald χ² statistic minus the degrees of freedom. Model discrimination was evaluated using the area under the ROC curve (AUC), calculated separately for women and men. Additional performance metrics, including accuracy, sensitivity, specificity, negative predictive value (NPV), and positive predictive value (PPV) were also reported for different score thresholds. Model calibration was evaluated using calibration plots and the Hosmer–Lemeshow goodness-of-fit test. To assess the incremental discrimination of the modified model over the H_2_FPEF score, we compared AUCs using the DeLong test(19). Risk reclassification was evaluated using the integrated discrimination improvement (IDI) and continuous and categorical net reclassification improvement (NRI)(20). Decision curve analysis was performed to compare the net clinical benefit of the models across a range of probability thresholds(21). As an internal validation, optimism corrected AUCs were estimated using bootstrap resampling with 500 iterations. In each bootstrap sample, models were refitted, and their performance was assessed both in the bootstrap sample and in the original dataset. The average difference between original and test performance represented the optimism, which was subtracted from the original AUC to obtain an optimism corrected estimate of discrimination. Findings are reported in accordance with the guidelines set forth in the TRIPOD statement. All tests are 2-sided, with a value of P<0.05 considered significant. All analyses were performed in R (version 4.4.1).

## RESULTS

### Baseline characteristics

A total of 515 patients with type 2 diabetes and aged 60 years or over were included from the UHFO-DM cohort, of whom 248 (48 %) were women (**Table 1**). Among the cohort, 123 patients (24 %) were diagnosed with HFpEF, 72 (29 %) women and 51 (19%) men. The overall mean age was 71 (± 7) years, with HFpEF patients being significantly older than non-HFpEF patients in both sexes. Specifically, women with HFpEF had a mean age of 75 years compared to 71 years in those without HFpEF, while men with HFpEF averaged 73 years versus 71 years in non-HFpEF men. Patients with HFpEF had a higher body mass index (BMI) in both sexes, and the proportion of individuals with obesity was markedly higher in the HFpEF patients in both women and men. Women with HFpEF presented significantly more often with a history of hypertension, while in men the difference did not reach statistical significance. Angina was more frequently reported in patients with HFpEF compared to non-HFpEF. In women with HFpEF, the prevalence of angina was 27.8% vs 10.2% in non-HFpEF and in men with HFpEF 35.3% vs 19.0% in non-HFpEF. The average number of antihypertensive drugs used were higher in the HFpEF patients than non-HFpEF patients in both sexes. NT-proBNP concentrations were significantly higher in HFpEF patients compared to non-HFpEF in both women and men.

**Table 1.** Baseline characteristics of all patients stratified by sex and HFpEF in the UHFO-DM cohort.

|  | All patients<br>(n=515) | Women<br>(n=248) |  |  | Men<br>(n=267) |  |  |
| --- | --- | --- | --- | --- | --- | --- | --- |
|  |  | Non-HFpEF<br>(n=176) | HFpEF<br>(n=72) | P value | Non-HFpEF<br>(n=216) | HFpEF<br>(n=51) | P value |
| Age, years | 71.4 (7.3) | 70.5 (7.2) | 75.0 (7.0) | <0.001 | 70.7 (6.8) | 73.1 (8.4) | 0.028 |
| BMI, kg/m<sup>2</sup> | 27.7 (4.2) | 27.5 (4.4) | 30.0 (5.1) | <0.001 | 26.8 (3.4) | 28.9 (3.8) | <0.001 |
| BMI>30 kg/m<sup>2</sup>, % | 134 (26.0) | 47 (26.7) | 34 (47.2) | 0.003 | 35 (16.2) | 18 (35.3) | 0.004 |
| Current smoker, % | 72 (14.0) | 24 (13.6) | 7 (9.7) | 0.526 | 29 (13.4) | 12 (23.5) | 0.113 |
| Angina, % | 97 (18.8) | 18 (10.2) | 20 (27.8) | 0.001 | 41 (19.0) | 18 (35.3) | 0.019 |
| Antihypertensive drugs, n | 1.20 (1.04) | 1.06 (1.01) | 1.58 (1.00) | <0.001 | 1.06 (1.02) | 1.69 (1.05) | <0.001 |
| Antihypertensive drugs≥2, % | 200 (38.8) | 61 (34.7) | 38 (52.8) | 0.012 | 72 (33.3) | 29 (56.9) | 0.003 |
| Comorbidities, % |  |  |  |  |  |  |  |
| Hypertension | 344 (66.8) | 116 (65.9) | 62 (86.1) | 0.002 | 128 (59.3) | 38 (74.5) | 0.063 |
| COPD | 19 (3.7) | 2 (1.1) | 2 (2.8) | 0.707 | 13 (6.0) | 2 (3.9) | 0.805 |
| Asthma | 42 (8.2) | 12 (6.8) | 14 (19.4) | 0.007 | 13 (6.0) | 3 (5.9) | 1.000 |
| Dyslipidemia | 364 (70.7) | 120 (68.2) | 52 (72.2) | 0.635 | 157 (72.7) | 35 (68.6) | 0.684 |
| Atrial fibrillation | 44 (8.5) | 8 (4.5) | 9 (12.5) | 0.048 | 16 (7.4) | 11 (21.6) | 0.006 |
| Valvular heart disease | 3 (0.6) | 1 (0.6) | 0 (0.0) | 1.000 | 1 (0.5) | 1 (2.0) | 0.831 |
| Cerebrovascular disease | 48 (9.3) | 13 (7.4) | 9 (12.5) | 0.299 | 18 (8.3) | 8 (15.7) | 0.183 |
| Prior MI/PCI/CABG | 55 (10.7) | 5 (2.8) | 8 (11.1) | 0.019 | 26 (12.0) | 16 (31.4) | 0.001 |
| Medication usage, % |  |  |  |  |  |  |  |
| ACEI/ARB | 177 (34.4) | 57 (32.4) | 21 (29.2) | 0.730 | 77 (35.6) | 22 (43.1) | 0.404 |
| Beta-blocker | 186 (36.1) | 59 (33.5) | 42 (58.3) | 0.001 | 57 (26.4) | 28 (54.9) | <0.001 |
| CCB | 84 (16.3) | 14 (8.0) | 18 (25.0) | 0.001 | 37 (17.1) | 15 (29.4) | 0.073 |
| Loop diuretics | 25 (4.9) | 7 (4.0) | 9 (12.5) | 0.028 | 2 (0.9) | 7 (13.7) | <0.001 |
| Thiazide diuretics | 128 (24.9) | 44 (25.0) | 18 (25.0) | 1.000 | 53 (24.5) | 13 (25.5) | 1.000 |
| Potassium-sparing diuretics | 17 (3.3) | 6 (3.4) | 6 (8.3) | 0.189 | 4 (1.9) | 1 (2.0) | 1.000 |
| Antiplatelet agents | 119 (23.1) | 25 (14.2) | 21 (29.2) | 0.010 | 57 (26.4) | 16 (31.4) | 0.587 |
| Anticoagulants | 22 (4.3) | 1 (0.6) | 5 (6.9) | 0.012 | 7 (3.2) | 9 (17.6) | <0.001 |
| Statins | 351 (68.2) | 117 (66.5) | 50 (69.4) | 0.762 | 147 (68.1) | 37 (72.5) | 0.649 |
| Oral antidiabetics | 336 (65.2) | 111 (63.1) | 50 (69.4) | 0.419 | 141 (65.3) | 34 (66.7) | 0.981 |
| Creatinine, $\mu\text{mol/l}$ | 79.7 (20.5) | 70.6 (15.2) | 70.1 (16.9) | 0.808 | 87.7 (21.1) | 90.2 (19.0) | 0.452 |
| CKD-EPI eGFR, $\mu\text{mol/l}$ | 67.6 (16.9) | 76.1 (15.4) | 74.5 (15.6) | 0.475 | 60.9 (14.7) | 57.3 (13.9) | 0.125 |
| NT-proBNP, pg/ml | 76.11<br>[42.29, 135.31] | 76.11<br>[42.29, 118.40] | 118.40<br>[67.66, 236.80] | <0.001 | 63.43<br>[33.83, 109.94] | 126.86<br>[71.88, 482.05] | <0.001 |
| E/e' ratio | 9.4 (3.4) | 9.5 (2.6) | 12.3 (6.1) | <0.001 | 8.3 (1.9) | 10.0 (3.1) | <0.001 |
| E/e' ratio>9, % | 238 (48.1) | 91 (52.6) | 53 (77.9) | 0.001 | 65 (31.0) | 29 (65.9) | <0.001 |
| PASP, mmHg | 14.0 (12.0) | 13.9 (11.9) | 15.1 (13.9) | 0.523 | 13.6 (11.0) | 14.7 (14.0) | 0.596 |
| PASP>35, % | 20 (4.5) | 8 (5.1) | 5 (8.5) | 0.550 | 5 (2.7) | 2 (5.0) | 0.788 |
| LVEF, % | 61.6 (6.8) | 62.1 (7.3) | 61.1 (6.6) | 0.271 | 62.0 (6.6) | 59.0 (5.3) | 0.002 |

ACEI, angiotensin-converting enzyme inhibitor; AF, atrial fibrillation; ARB, angiotensin receptor blocker; BMI, body mass index; CABG, coronary artery bypass grafting; CCB, calcium channel blocker; CI, confidence interval; CKD-EPI, Chronic Kidney Disease Epidemiology Collaboration; COPD, chronic obstructive pulmonary disease; eGFR, estimated glomerular filtration rate; HFpEF, heart failure with preserved ejection fraction; LVEF, left ventricular ejection fraction; MI, myocardial infarction; NT-proBNP, N-terminal pro-B-type natriuretic peptide; PASP, pulmonary artery systolic pressure; PCI, percutaneous coronary intervention.

Creatinine levels were slightly higher and eGFR tended to be lower in the HFpEF patients, although differences did not reach statistical significance. Although LVEF remained within the preserved range (above 50%) in HFpEF and non-HFpEF patients, patients with HFpEF, particularly men, exhibited lower LVEF values.

### H_2_FPEF score overall and sex-specific performance

In the UHFO-DM cohort, the H_2_FPEF score demonstrated moderate discriminatory ability for identifying HFpEF, with an AUC of 0.72 (95% CI: 0.67–0.77) (**Figure 1, Supplementary Table 2**). Sensitivity and specificity were 0.69 and 0.64, respectively, with a PPV of 0.37 and an NPV of 0.87. In addition, discrimination by AUC was higher in men (0.74, 95% CI: 0.66–0.81) than in women (0.69, 95% CI: 0.62–0.76), although the unpaired Delong test did not reach statistical significance. For other diagnostic characteristics, sensitivity in women was only 0.53 compared to 0.69 in men, and NPV was also lower in women (0.80) than in men (0.91). The calibration plot by score showed decent calibration in overall, women and men population (**Supplementary Figure 2**).

**Figure 1.**
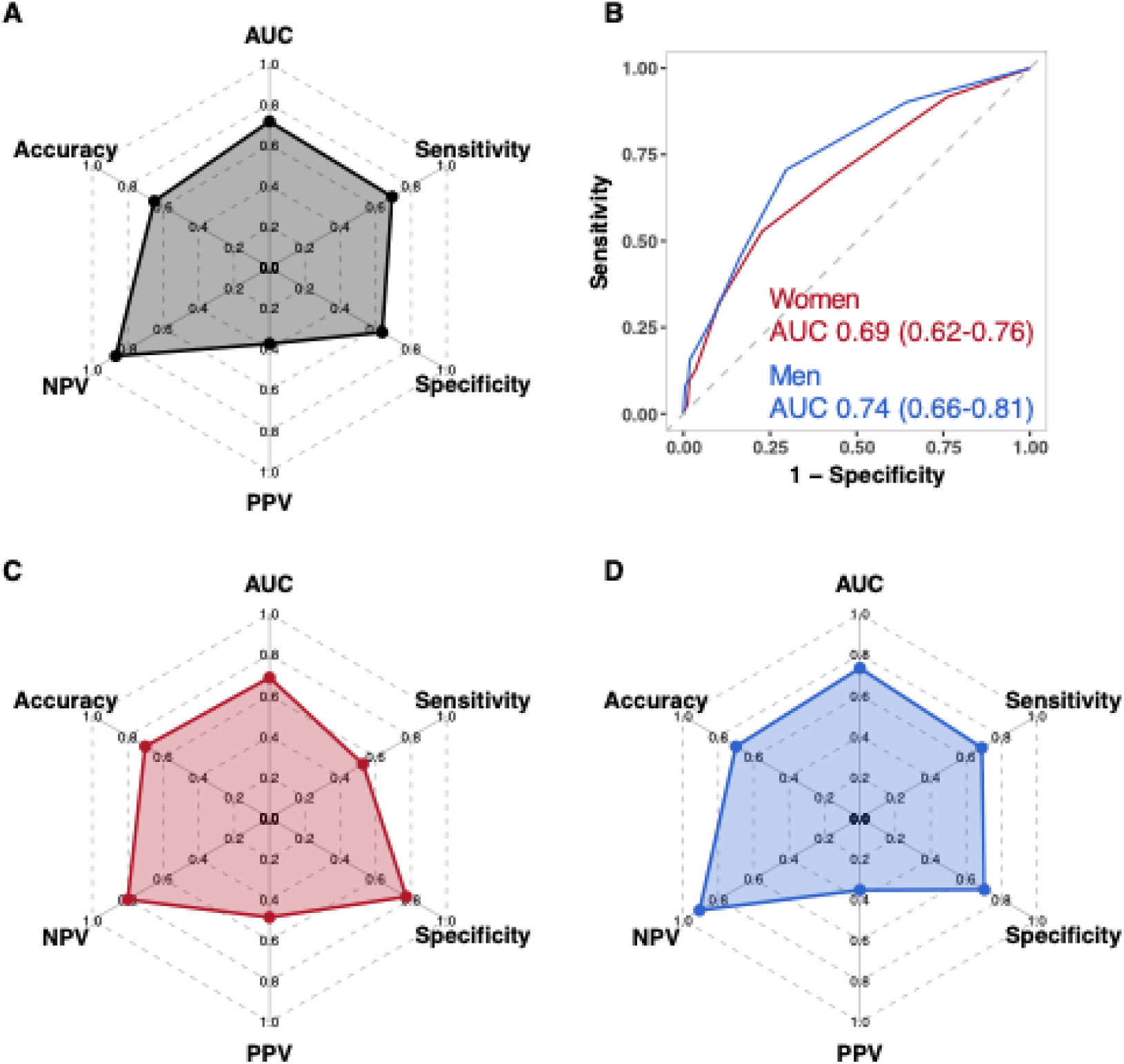
Diagnostic performance of the H_2_FPEF score in the UHFO-DM cohort. Radar plots show accuracy, sensitivity, specificity, PPV, NPV, and AUC for the overall population (A), women (C), and men (D). ROC curves are presented separately for women and men (B). AUC, area under the curve; HFpEF, heart failure with preserved ejection fraction; NPV, negative predictive value; PPV, positive predictive value; ROC, receiver operating characteristic.

### Association between angina and HFpEF

In the UHFO-DM cohort, angina was associated with HFpEF in both women (OR 3.38, 95% CI: 1.66–6.86) and men (OR 2.33, 95% CI: 1.19– 4.54) in the univariable analyses (**Table 2**). When all six components of the H_2_FPEF score were included in multivariable models alongside angina, angina remained independently associated with HFpEF in women (OR 3.96, 95% CI: 1.72–9.11), but not in men (OR 1.90, 95% CI: 0.88–4.10). In the univariable analysis, five individual components of the H_2_FPEF score were significantly associated with HFpEF in both women and men (all P < 0.05), except for PASP, which showed no association in either sex. In the multivariable analysis, age, BMI, and E/e′ ratio remained significant predictors in both sexes, while atrial fibrillation and PASP were not independently associated with HFpEF (antihypertensives significant in men but not in women). Continuous variables were standardized using z-scores to allow direct comparison of effect sizes in the forest plots (**Figure 2**). Regarding feature importance in the logistic model, based on Wald χ² minus degrees of freedom, angina ranked as the third most informative variable in women, after BMI and age, but had lower relative importance in men. To further confirm the incremental predictive value of angina beyond the composite H_2_FPEF score, we constructed additional multivariable models using the total H_2_FPEF score as a single covariate. In women, angina remained independently significantly associated with HFpEF after adjustment for the total score (OR 3.21, 95% CI: 1.51–6.81; P = 0.003). In contrast, in men, angina was not independently associated with HFpEF (OR 1.67, 95% CI: 0.81–3.45; P = 0.168).

**Figure 2.**
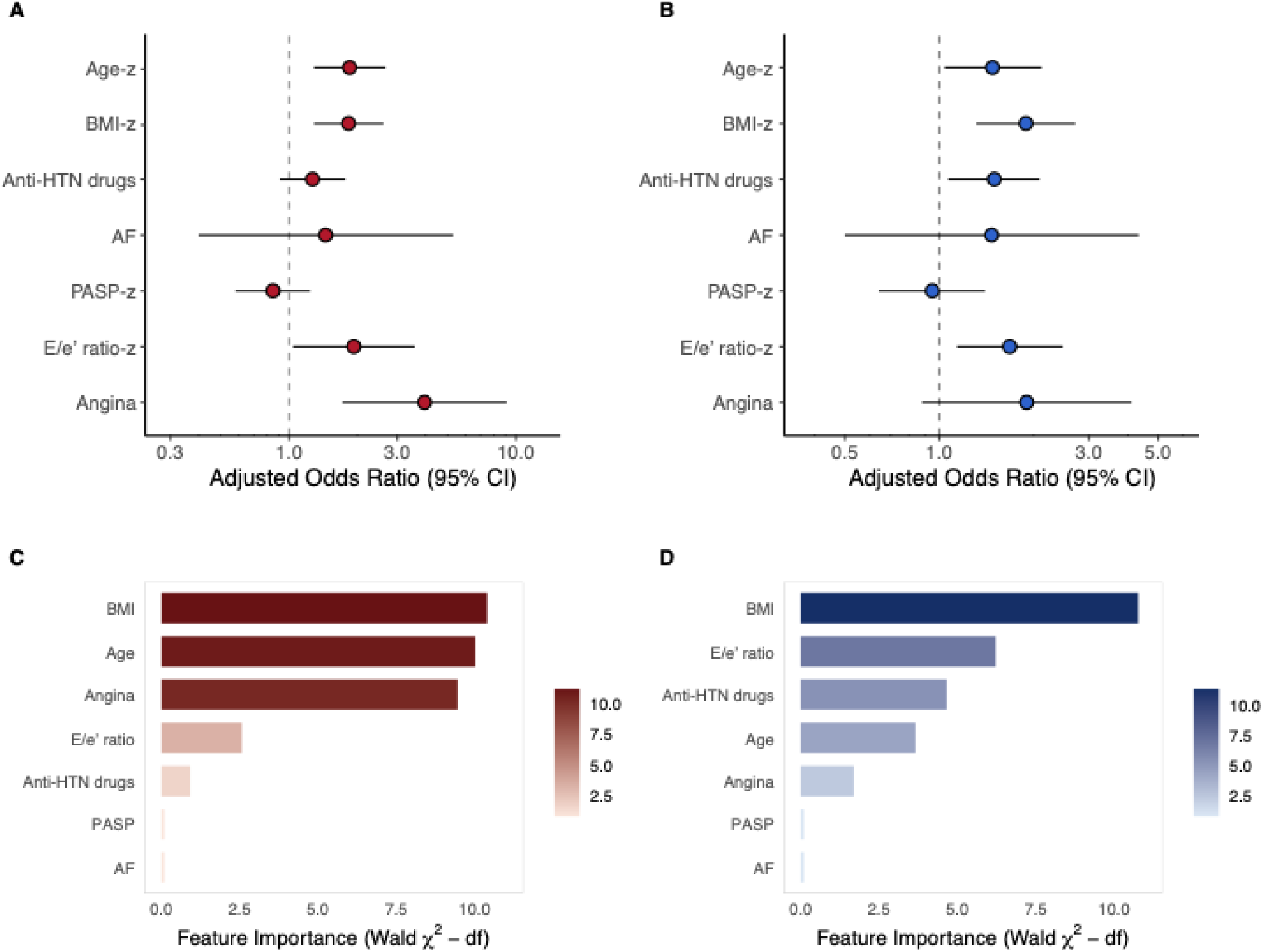
Multivariable analysis of H_2_FPEF components and angina in the UHFO-DM cohort. Forest plots display odds ratios with 95% CI for H_2_FPEF components and angina in women (A) and men (B). Variables with “-z” were standardized as z-scores to allow comparability. Variable importance, quantified by Wald χ² minus degrees of freedom, is shown in bar plots for women (C) and men (D). AF, atrial fibrillation; Anti-HTN drugs, antihypertensive drugs; BMI, body mass index; CI, confidence interval; PASP, pulmonary artery systolic pressure.

**Table 2.** Univariable and multivariable logistic regression of H_2_FPEF and angina with HFpEF in the UHFO-DM cohort.

|  | Women |  |  |  | Men |  |  |  |
| --- | --- | --- | --- | --- | --- | --- | --- | --- |
|  | Univariable |  | Multivariable<br>(Factors and angina) |  | Univariable |  | Multivariable<br>(Factors and angina) |  |
|  | OR (95%CI) | P value | OR (95%CI) | P value | OR (95%CI) | P value | OR (95%CI) | P value |
| Age, years | 1.09 (1.05–1.13) | <0.001 | 1.09 (1.03–1.14)<br>1.85 (1.29–2.67) * | 0.001 | 1.05 (1.00–1.09) | 0.031 | 1.06 (1.01–1.11)<br>1.48 (1.04–2.12) * | 0.032 |
| BMI, kg/m<sup>2</sup> | 1.11 (1.05–1.18) | 0.001 | 1.14 (1.06–1.22)<br>1.83 (1.29–2.61) * | 0.001 | 1.16 (1.07–1.27) | 0.001 | 1.20 (1.08–1.33)<br>1.89 (1.31–2.72) * | 0.001 |
| Antihypertensive drugs, n | 1.64 (1.24–2.15) | 0.001 | 1.27 (0.91–1.77) | 0.168 | 1.73 (1.29–2.32) | <0.001 | 1.50 (1.07–2.09) | 0.018 |
| Atrial fibrillation | 3.00 (1.11–8.12) | 0.032 | 1.45 (0.40–5.29) | 0.577 | 3.44 (1.48–7.96) | 0.004 | 1.47 (0.50–4.33) | 0.481 |
| E/e' ratio | 1.19 (1.06–1.35) | 0.008 | 1.14 (1.00–1.30)<br>1.93 (1.04–3.59) * | 0.044 | 1.34 (1.17–1.54) | 0.000 | 1.24 (1.06–1.45)<br>1.68 (1.14–2.48) * | 0.008 |
| Pulmonary artery systolic pressure, mmHg | 1.01 (0.98–1.03) | 0.584 | 0.99 (0.96–1.02)<br>0.85 (0.58–1.24) * | 0.395 | 1.01 (0.98–1.04) | 0.387 | 1.00 (0.96–1.03)<br>0.95 (0.64–1.40) * | 0.786 |
| Angina | 3.38 (1.66–6.86) | 0.001 | 3.96 (1.72–9.11) | 0.001 | 2.33 (1.19–4.54) | 0.014 | 1.90 (0.88–4.10) | 0.102 |
|  | Univariable |  | Multivariable<br>(Score and angina) |  | Univariable |  | Multivariable<br>(Score and angina) |  |
| Score | 1.54 (1.28–1.85) | <0.001 | 1.52 (1.27–1.83) | <0.001 | 1.67 (1.37–2.03) | <0.001 | 1.62 (1.33–1.98) | <0.001 |
| Angina | 3.38 (1.66–6.86) | 0.001 | 3.21 (1.51–6.81) | 0.003 | 2.33 (1.19–4.54) | 0.014 | 1.67 (0.81–3.45) | 0.168 |

The associations between HFpEF and angina, components of H<sub>2</sub>FPEF and H<sub>2</sub>FPEF score are assessed by logistic regression. Standardized variables are marked with an asterisk.

### Development, evaluation and internal validation of the modified H_2_FPEF score

Above findings provided a rationale for incorporating angina into the H_2_FPEF score in women. Angina remained significantly associated with HFpEF after adjusting for the total score (β estimate for angina: 1.17; for H_2_FPEF score: 0.42). According to the H_2_FPEF score construction principles, where point values were assigned based on effect sizes from logistic regression, this would theoretically translate into a weight of approximately 2.8 for angina, suggesting a possible allocation of 3 points. However, given the potential subjectivity and variability in the clinical interpretation of patient-reported angina symptoms, we aimed to avoid disproportionately amplifying its influence on the overall score. To preserve model stability and retain clinical interpretability, we conservatively assigned angina a weight of 1 point in the modified H_2_FPEF score. A version assigning 3 points to angina was also tested and is presented in the **Supplementary Figure 3**. The modified H_2_FPEF model demonstrated superior discrimination for HFpEF compared with the original H_2_FPEF score (**Figure 3**). The AUC increased from 0.69 (95% CI: 0.62–0.76) to 0.71 (95% CI: 0.64–0.78), with a statistically significant difference by DeLong test (P = 0.030). Internal validation with 500 bootstrap iterations showed negligible optimism, with optimism-corrected AUCs of 0.69 for the original H_2_FPEF model and 0.71 for the modified model, confirming stable discriminatory performance after adjustment for optimism (**Supplementary Figure 4**). The modified model also achieved a lower AIC (273.1 vs. 278.7), indicating better overall model fit. The modified score improved sensitivity (0.60 vs. 0.53), PPV (0.51 vs. 0.49), NPV (0.82 vs. 0.80), and overall accuracy (0.71 vs. 0.70), while maintaining similar specificity (0.76 vs. 0.77) compared to the original score (**Supplementary Table 3**). Radar plots visualized incremental gain of these metrics in overall diagnostic performance. The modified model also showed decent calibration. The Hosmer–Lemeshow goodness-of-fit test indicated an adequate model fit (P = 0.373), and visual inspection of the calibration curve indicated close alignment between predicted probabilities and observed prevalence of HFpEF across risk strata (**Supplementary Figure 5**). Risk reclassification analysis confirmed the added value of the modified score. Compared with the original model, the modified H_2_FPEF score yielded a continuous NRI of 0.449 (95% CI: 0.199–0.710; P = 0.001) and an IDI of 0.023 (95% CI: 0.012–0.034; P < 0.001), indicating its superior performance as regards risk reclassification. The categorical reclassification tables are presented in **Supplementary Table 4**, showing that the modified model had better rule-out performance compared with the original model. Decision curve analysis demonstrated higher net clinical benefit for the modified score across a wide range of threshold probabilities, further supporting its utility in clinical decision-making for women with suspected HFpEF. The modified H_2_FPEF score in women (heavy, 2 or more hypertensive drugs, atrial fibrillation, pulmonary hypertension, elder age>60, elevated filling pressures, angina) ranges from 0 to 10, which is illustrated in the nomogram (**Figure 4**). The probability of HFpEF increases with increasing modified H_2_FPEF score, and the score-to-probability conversion table is provided in **Supplementary Table 5**. In men, the association between angina and HFpEF was not statistically significant. Nevertheless, for robustness, we also constructed a modified H_2_FPEF score for men in the same approach with women. The results showed no improvement in diagnostic performance: the AUC changed from 0.74 (95% CI: 0.66–0.81) to 0.73 (95% CI: 0.67–0.81), with no significant difference by the DeLong test (P = 0.769; **Supplementary Figure 6**). This confirms that angina does not add diagnostic value for HFpEF in men.

**Figure 3.**
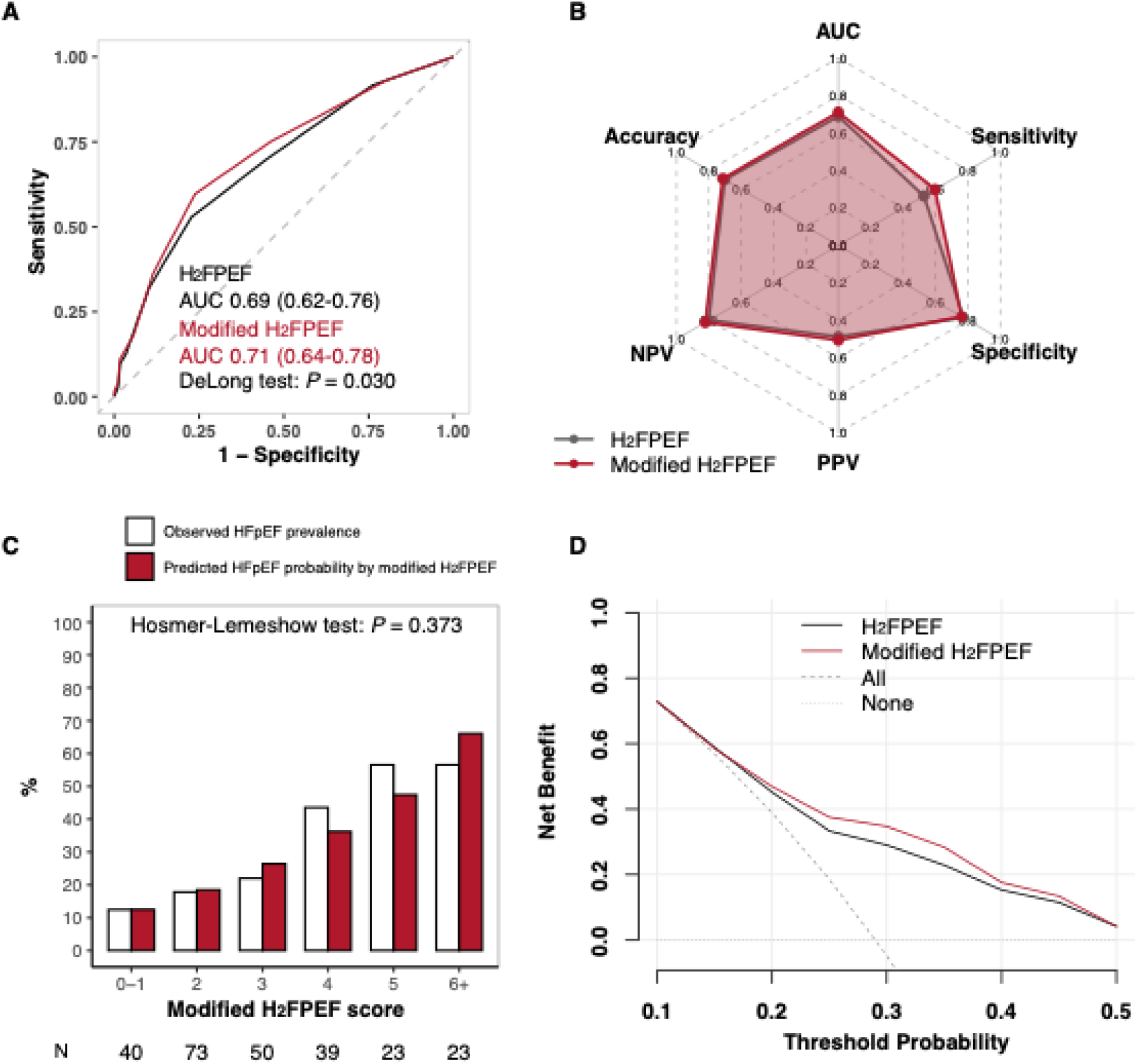
Comparison of the original and modified H_2_FPEF score in women of the UHFO-DM cohort. Panel A shows ROC curves for the original and modified H_2_FPEF score. Panel B displays radar plots of accuracy, sensitivity, specificity, PPV, NPV, and AUC. Panel C shows calibration with observed versus predicted probabilities across categories of the modified score. Panel D presents decision curve analysis comparing net clinical benefit across a range of threshold probabilities. AUC, area under the curve; HFpEF, heart failure with preserved ejection fraction; NPV, negative predictive value; PPV, positive predictive value; ROC, receiver operating characteristic.

**Figure 4.**
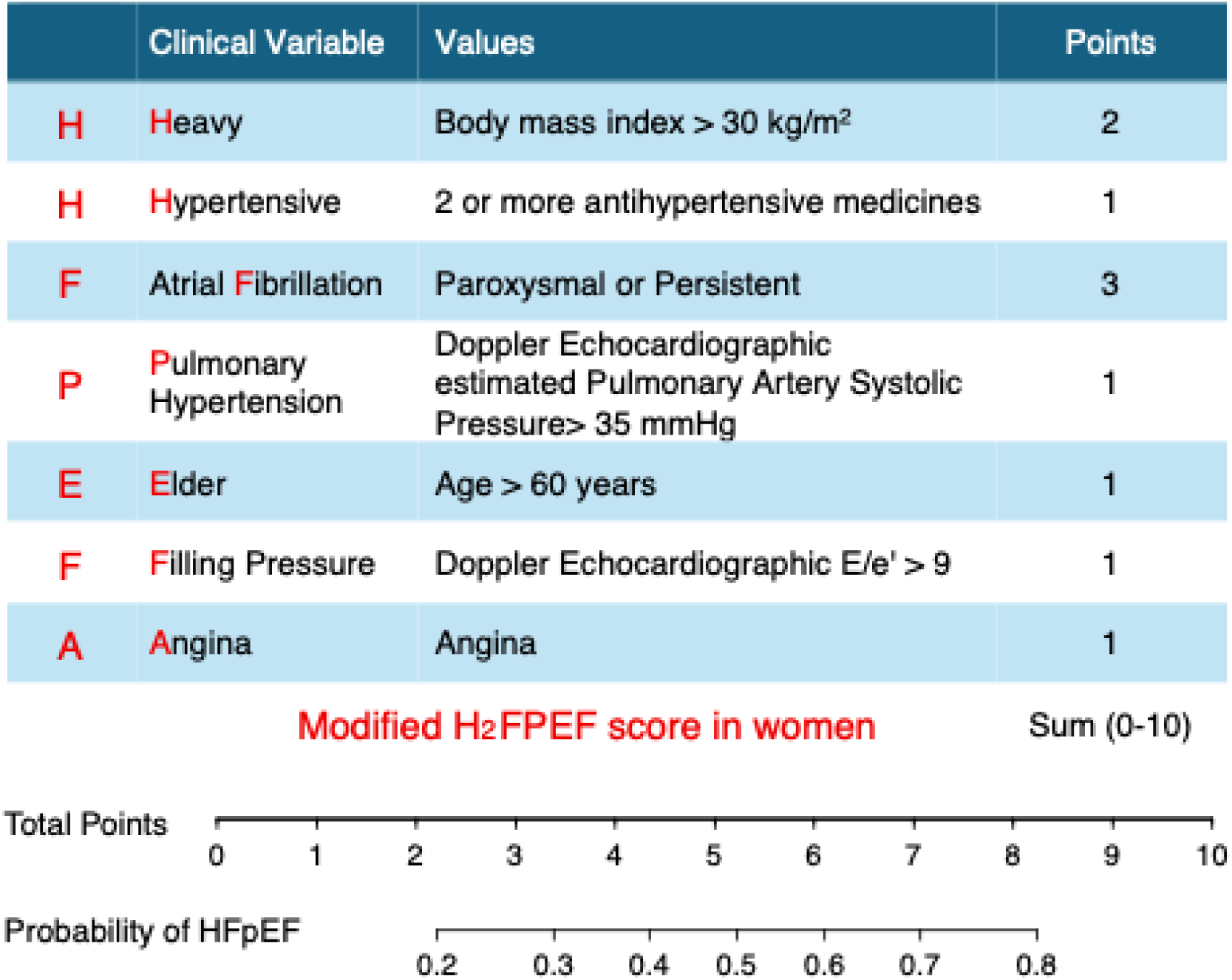
Modified H_2_FPEF score in women. The table lists clinical variables, definitions, and assigned points for the modified H_2_FPEF score in women. The total score ranges from 0 to 10. The probability of HFpEF corresponding to each score is shown below. AF, atrial fibrillation; BMI, body mass index; HFpEF, heart failure with preserved ejection fraction; PASP, pulmonary artery systolic pressure.

### External validation of association between angina and HFpEF

In the combination cohort (**Supplementary Table 6**), univariable logistic regression showed that angina was significantly associated with HFpEF in women (OR 1.91, 95% CI: 1.13–3.22, *P* = 0.016), but not in men (OR 1.09, 95% CI: 0.61–1.97, *P* = 0.768) (**Supplementary Table 7**). Due to extensive missingness of PASP, this variable was not included in the multivariable models. When adjusted for the other five individual H_2_FPEF score components, angina remained independently associated with HFpEF in women (OR 2.13, 95% CI: 1.14–3.97, *P* = 0.018), but not in men (OR 0.85, 95% CI: 0.44–1.66, *P* = 0.638) (**Figure 5**).

**Figure 5.**
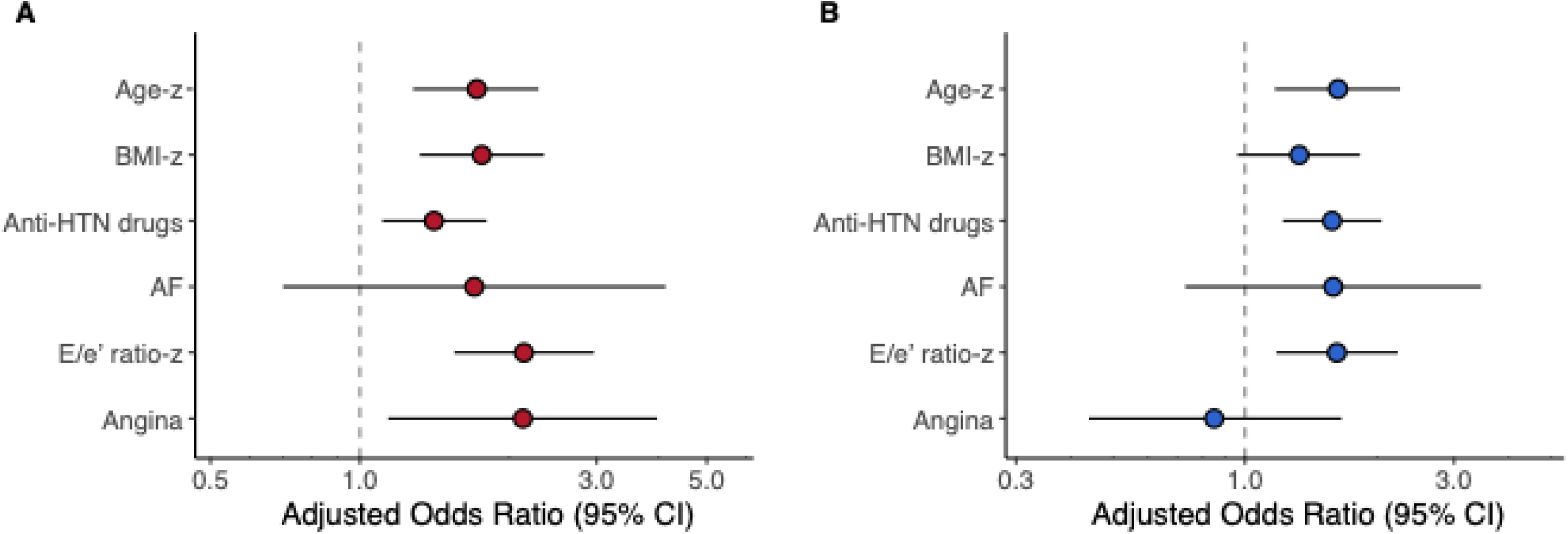
External validation of association between angina and HFpEF. Forest plots display odds ratios with 95% CI for H_2_FPEF components, including angina in women (A) and men (B). Variables with “-z” were standardized as z-scores to allow comparability. AF, atrial fibrillation; Anti-HTN drugs, antihypertensive drugs; BMI, body mass index; CI, confidence interval.

## Discussion

In this study, we observed that the H_2_FPEF score had moderate diagnostic performance for HFpEF in our population (AUC 0.72), but the sex-specific performance is limited (0.74 in men and 0.69 in women), and the lower sensitivity in women may limit its ability to reliably detect HFpEF in women. On the other hand, angina was independently associated with HFpEF in women but not in men after adjusting for the total H_2_FPEF score or its individual components. In women, the inclusion of angina in a modified H2PEF score improved diagnostic performance, increasing the AUC from 0.69 to 0.71 and sensitivity from 0.53 to 0.60, while preserving good calibration. The modified H_2_FPEF score also significantly improved risk reclassification, underscoring the added value of angina for HFpEF diagnosis in women.

Angina is a common clinical manifestation in HFpEF, however, not very well appreciated. Robert et al. reported that approximately 40% of HFpEF patients from the Duke Databank for Cardiovascular Disease experienced recent angina(12), which is broadly consistent with the 30% prevalence observed in our cohorts. In a prospective cohort of hospitalized HFpEF patients, approximately 51% were found to have obstructive CAD(22), which may partially explain angina symptoms in HFpEF patients. However, obstructive CAD alone does not fully account for the burden of angina in HFpEF. Non-obstructive, especially CMD, has emerged as another key mechanism. This is supported by a meta-analysis of over 800 HFpEF patients, which reported an overall CMD prevalence of approximately 71%(23). These findings suggest that microvascular ischemia, independent of overt epicardial coronary obstruction, may also be an important driver of angina in HFpEF patients. The mechanistic interplay among HFpEF, CMD, and angina has been increasingly recognized. HFpEF and microvascular angina are increasingly recognized as distinct but related syndromes that share a common basis in coronary microvascular pathology(24). This perspective contributes to an evolving paradigm in which HFpEF is no longer viewed solely as the potential consequence of hypertension and ventricular stiffening, but rather as a systemic inflammatory syndrome which also involve the coronary microcirculation(25).

Importantly, the relationships between HFpEF, obstructive CAD, and CMD appear to be sex specific. In a retrospective study of 376 HFpEF patients, those with obstructive CAD were significantly more likely to be male compared to those without CAD (57% vs. 25%)(26). Similarly, in the PARAMOUNT study, women with HFpEF had a lower prevalence of prior MI compared to men with HFpEF (11% vs. 22%)(27). Comparable patterns were seen in our data, with prior MI/PCI/CABG prevalence rates of 11.1% vs. 31.4% in the UHFO-DM cohort and 9.6% vs. 25.8% in the combination cohort. In contrast, women with HFpEF are more likely to present with non-obstructive CAD and underlying CMD (28, 29). Longitudinal data from the Women’s Ischemia Syndrome Evaluation (WISE) study further demonstrated that women with angina symptoms but no obstructive CAD were at increased risk of subsequently developing HFpEF(30). Taken together, although the sex-specific mechanisms of angina in HFpEF require further investigation, they may play an important role in the sex-related association between angina and HFpEF, supporting a sex-stratified approach to modifying diagnostic tools.

Our work was motivated by the observation that conventional HFpEF diagnostic algorithms, while effective in many contexts, may be less sensitive to certain female-specific phenotypes. The original H_2_FPEF score has been widely validated as a practical and accessible tool for identifying HFpEF across diverse clinical settings(31, 32). However, it does not take sex differences into account, despite the fact that HFpEF disproportionately affects women(33), and female patients often present with distinct clinical patterns, including differences in risk factor profiles and outcomes(25). Notably, women comprised the majority (61%) of the HFpEF population in the score’s derivation cohort(8).

Nonetheless, sex was not retained as an independent predictive variable, possibly because the control group with non-cardiac dyspnea also included a similar proportion of women (59%). This could partly explain why the score’s sensitivity is lower in women in our study, particularly those with atypical features, such as non-obese patients whose HFpEF may be driven by mechanisms like microvascular ischemia and who might therefore have lower H_2_FPEF scores despite having the condition. To address this, our modified H_2_FPEF score introduced angina pectoris as an additional variable specifically in women. This addition resulted in improved diagnostic performance in women: sensitivity increased, and the negative predictive value also improved, reinforcing its utility as a rule-out tool. Clinically, this implies that a woman without angina and with a low modified score is unlikely to have HFpEF, whereas a woman with borderline features but presenting with angina warrants further diagnostic evaluation.

From a clinical standpoint, the modified H_2_FPEF score we propose for women remains consistent with the original score’s intent, which is simple and composed entirely of easily obtainable clinical variables. Angina pectoris can be assessed through a single history question, and its inclusion does not add burden to patients and clinicians operating in time-constrained environments. This is a key advantage for a diagnostic tool intended for widespread use in evaluating unexplained dyspnea or suspected HFpEF cases. Moreover, this sex-tailored approach aligns with the growing emphasis in cardiovascular medicine on the recognition of sex-specific disease manifestations, thereby promoting diagnostic equity and improving sensitivity in populations that may otherwise be underrecognized(34). We acknowledge several limitations in our study. First, all participants in our cohorts were older than 60 years, which limited the generalizability of our findings to younger people. Moreover, everybody in our study had a value of at least one on the H_2_FPEF score. Second, the insignificant findings in men should be interpreted with caution given the moderate sample size, warranting further validation in larger datasets. Third, angina was assessed through patient self-report, which may introduce subjective variability. Therefore, we assigned it a relatively conservative weight in the modified model; nevertheless, its inclusion still led to a significant improvement in diagnostic performance. Fourth, the derivation cohort consisted exclusively of individuals with type 2 diabetes, whereas the validation cohort represented a broader population. However, because PASP was largely missing in the validation cohort, we were only able to confirm that angina retained independent predictive value for HFpEF in women after adjustment for the H_2_FPEF components, but could not externally validate the modified score itself. Future studies with complete echocardiographic data are warranted to address this limitation. Finally, our study lacked invasive hemodynamic testing as a diagnostic standard for HFpEF. However, it is worth noting that even invasive measures are not universally unified due to variations in exercise or loading protocols and threshold definitions. Expert panel diagnosis remains a widely adopted and reasonably reliable diagnostic approach in clinical practice (31, 32).

## Conclusion

In conclusion, our study demonstrated that incorporating presence of angina symptoms as an additional variable into the H_2_FPEF score improves the diagnostic performance of HFpEF in women. This modification enhances the model’s discrimination, sensitivity and rule-out capability in women, offering a more tailored and inclusive approach to HFpEF diagnosis. As clinical practice moves toward precision medicine, our findings support the integration of sex-specific clinical markers into established diagnostic tools to better serve diverse patient populations.

## Declarations

### Ethics approval and consent to participate

All used cohort studies were approved by the Medical Ethics Committee of the University Medical Center Utrecht, the Netherlands. All participants provided written informed consent.

## Availability of data and materials

The datasets used and analysed during the current study are available from the corresponding author on reasonable request.

## Authors’ contributions

E.C. and H.R. contributed to the conception and design of the work. K.Q., NC.OM., A.V., AM.O., Y.M., E.R., L.BW., M.H., M.C., A.T., R.M., and F.R. contributed to the acquisition, analysis, or interpretation of data for the work. K.Q. drafted the manuscript. E.C and H.R. critically revised the manuscript for important intellectual content. K.Q. performed the statistical analysis. H.R. obtained funding. NC.OM., and H.R. and E.C. supervised the work. All authors gave final approval and agreed to be accountable for all aspects of the work, ensuring integrity and accuracy.

### Abbreviations

ACC: American college of cardiology
ACEI: Angiotensin-converting enzyme inhibitor
AF: Atrial fibrillation
AIC: Akaike information criterion
ARB: Angiotensin receptor blocker
AUC: Area under the (ROC) curve
BMI: Body mass index
CABG: Coronary artery bypass grafting
CAD: Coronary artery disease
CCB: Calcium channel blocker
CI: Confidence interval
CKD-EPI: Chronic kidney disease epidemiology collaboration
CMD: Coronary microvascular dysfunction
COPD: Chronic obstructive pulmonary disease
ECG: Electrocardiography
eGFR: Estimated glomerular filtration rate
E/e′: Ratio of early mitral inflow velocity to mitral annular early diastolic velocity
ESC: European society of cardiology
HFpEF: Heart failure with preserved ejection fraction
HFrEF: Heart failure with reduced ejection fraction
IQR: Interquartile range
LVEF: Left ventricular ejection fraction
MI: Myocardial infarction
MICE: Multiple imputation by chained equations
NPV: Negative predictive value
NT-proBNP: N-terminal pro–B-type natriuretic peptide
NRI: Net reclassification improvement
OR: Odds ratio
PASP: Pulmonary artery systolic pressure
PCI: Percutaneous coronary intervention
PPV: Positive predictive value
ROC: Receiver operating characteristic
SD: Standard deviation
TDI: Tissue Doppler imaging
VIF: Variance inflation factor

## Acknowledgements

Language improvement in this manuscript was assisted by ChatGPT, in accordance with ethical guidelines for use and proper acknowledgment of generative AI in academic research.

